# Use of electronic health records to develop an actionable taxonomy of persistent hypertension

**DOI:** 10.1101/2022.11.01.22281573

**Authors:** Yuan Lu, Cindy Xinxin, Hazar Khidir, César Caraballo, Shiwani Mahajan, Erica S. Spatz, Leslie Curry, Harlan M. Krumholz

## Abstract

**Background:** The digital transformation of medical data presents opportunities for novel approaches to manage patients with persistent hypertension (defined as multiple measurements of elevated BP over 6 months). We sought to develop an actionable taxonomy of patients with persistent hypertension based on clinical data from the electronic health records (EHR).

**Methods:** This qualitative study was a content analysis of clinician notes in the EHR of patients in the Yale New Haven Health System. Eligible patients were 18 to 85 years and had blood pressure ≥160/100 mmHg at five or more consecutive outpatient visits between January 1^st^ 2013 to October 31^st^ 2018. A total of 4,828 patients met criteria, of which 200 records were randomly selected for chart review. Through a systematic, inductive approach, we developed a rubric to abstract data from the EHR and then analyzed the abstracted data qualitatively using conventional content analysis until saturation was reached.

**Results:** We reached saturation with 115 patients, who had a mean age of 68.1 (SD, 11.6) years; 54.8% were female; 52.2%, 30.4%, and 13.9% were White, Black, and Hispanic people. We identified three content domains related to persistence of hypertension: (1) non-intensification of pharmacological treatment (defined as absence of antihypertensive treatment intensification in response to persistent severely elevated blood pressure) with four subcategories, including provider purview, competing medical priorities, patient preference, and de-emphasis of the office measurement; (2) non-implementation of prescribed treatment (defined as a documentation of provider recommending a specified treatment plan to address hypertension but treatment plan not being implemented) with four subcategories, including obstacles to obtaining medications, psychosocial barriers, patient misunderstanding, and negative medication experience; and (3) non-response to prescribed treatment (defined as clinician-acknowledged persistent hypertension despite documented effort to escalate existing pharmacologic agents and addition of additional pharmacologic agents with presumption of adherence) with two subcategories, including resistant hypertension and secondary hypertension.

**Conclusions:** This study presents a novel actionable taxonomy for classifying patients with persistent hypertension by their contributing causes based on EHR data. These categories can be automated and linked to specific types of actions to address them.

**Clinical Perspective:** *What is Known?:*

- This study developed a novel actionable taxonomy for classifying patients with persistent hypertension by their contributing causes, using qualitatively content analysis of clinician notes in the EHR.
- We identified three main content domains and a variety of subcategories contributing to persistent hypertension (non-intensification of pharmacological treatment, non-implementation of prescribed treatment, non-response to prescribed treatment), providing actionable information to inform solutions.
- This taxonomy is based on real-world data in the EHR, so it is pragmatic for use in the clinical setting.

*What the Study Adds?:*

- This actionable taxonomy lays the foundation for developing effective tools for health systems to rapidly identify and classify people with persistent hypertension and connect them with targeted, personalized interventions at scale.
- Personalized medicine depends on distinguishing patients with persistent hypertension by their contributing factors; such knowledge is essential for tailoring care to individuals with appropriate evidence-based interventions.

## BACKGROUND

Despite the availability of effective therapy, the United States has made little progress in controlling hypertension. In the past 20 years, the percentage of hypertensive adults who achieve the blood pressure (BP) control (defined as BP <140/90 mmHg) increased from 32% in 1999-2000 to 54% in 2013-2014, and then dropped to 44% in 2017-2018.^1^ This percentage further dropped to 19% using the 2017 American College of Cardiology (ACC)/American Heart Association (AHA) guideline BP control definition (BP <130/80 mmHg).^1, 2^ Thus, hypertension remains a leading cause of preventable deaths in the country.^3, 4^

Currently, many people have persistent hypertension, defined as multiple measurements of elevated BP over time (e.g., 6 months).^5^ These people need more support than they currently receive to achieve BP goals. Persistent hypertension is a more encompassing concept than resistance hypertension, which focuses on patients who have elevated BP despite the concurrent use of three antihypertensive drug classes at maximally tolerated doses.^6^ Patients with persistent hypertension may include people with absence of a diagnosis, inadequate treatment or lifestyle modification, poor adherence, missed appointments, resistant hypertension, and those with other barriers to health care. Given the many reasons why patients may have persistent hypertension and the different strategies to address these reasons, it is critical to have a better, more nuanced way to classify these patients so that we can provide interventions targeted to the obstacles to their BP goals.

The electronic health records (EHR) provide efficient access to a wide range of longitudinal clinical information that can be used to better identify and classify patients with persistent hypertension so as to facilitate personalized interventions.^7, 8^ Yet, to date, this promise is unfulfilled as no standard way is available to classify patients with persistent hypertension. A pragmatic classification system that can leverage EHR data is needed to provide actionable information to guide the selection of intervention strategies, with particular attention to problems related to implementation of effective antihypertensive strategies.

Accordingly, we used qualitative content analysis of the EHR to develop a pragmatic taxonomy that distinguishes patients with persistent severe hypertension by their contributing factors, with a focus on the actionability of the classification.^9, 10^ We focused on people with severe hypertension (defined as multiple measurements of BP>=160/100 mmHg over time) with the presumption that the signal for impediments would be strongest in this group, and they were people at the highest risk of adverse outcomes and with the most important need for intervention. Our thesis was that the impediments to effective BP control in these patients likely spans domains, including social determinants, provider-, patient-, and system-level barriers, and there was a need to be particularly attentive to factors undermining the implementation of effective strategies. We posited using an implementation science orientation that a useful classification could leverage EHR data to illuminate how provider-, patient-, and system-level factors contribute to persistent hypertension. Insights from this analysis could enable better classification of patients by their contributing factors to persistent hypertension and connect that classification to personalized interventions matching the impediments and enhancing outcome.^11^

## METHODS

### Data Sources

All data produced in the present work are contained in the manuscript. Data was drawn from an existing cohort of adult patients with at least two outpatient visits between January 1^st^, 2013 and October 31^st^, 2018 from Yale New Haven Health System (YNHHS). YNHHS is a large academic health system consisting of five distinct hospital delivery networks and associated ambulatory clinics located in Connecticut and Rhode Island. The system provides services for approximately two million patients annually. All YNHHS hospitals used a secure, centralized EHR system that is designed by the Epic Corporations to collect and store clinical and administrative claims data. The EHR data are maintained in a data repository at the YNHHS sever. This study was approved by the institutional review board at Yale University and informed consent was waived.

### Study Population

Eligible patients were 18 to 85 years old and had persistent severe hypertension, defined as having measurements of systolic BP (SBP) ≥160 mmHg or diastolic BP (DBP) ≥100 mmHg at five or more consecutive outpatient visits between January 1^st^, 2013 and October 31^st^, 2018. We required each visit to be at least 24 hours apart and the duration of five visits to be at least 30 days. We included only BP measurements recorded at ambulatory visits. We excluded BP measurements from inpatient and emergency department (ED) encounters because patients may have transiently elevated BP from acute medical conditions. We also excluded patients who were pregnant or on dialysis (**Figure 1**). We chose to focus on patients with severe hypertension as they are a priority population and the signals of impediments to achieving BP control are likely to be strongest. We required any two consecutive visits to be at least one day apart. We had access to all available data in the medical records, including patient demographics, past medical histories, vital signs, outpatient medications, laboratory results, encounter notes and scanned documents. A total of 4,828 patients met the eligibility criteria. We randomly selected 200 records from the group of all eligible patients for qualitative analysis, with the intent to select more if we did not achieve saturation (where no new concepts emerge from analyses of subsequent data).^12^

**Figure 1.**
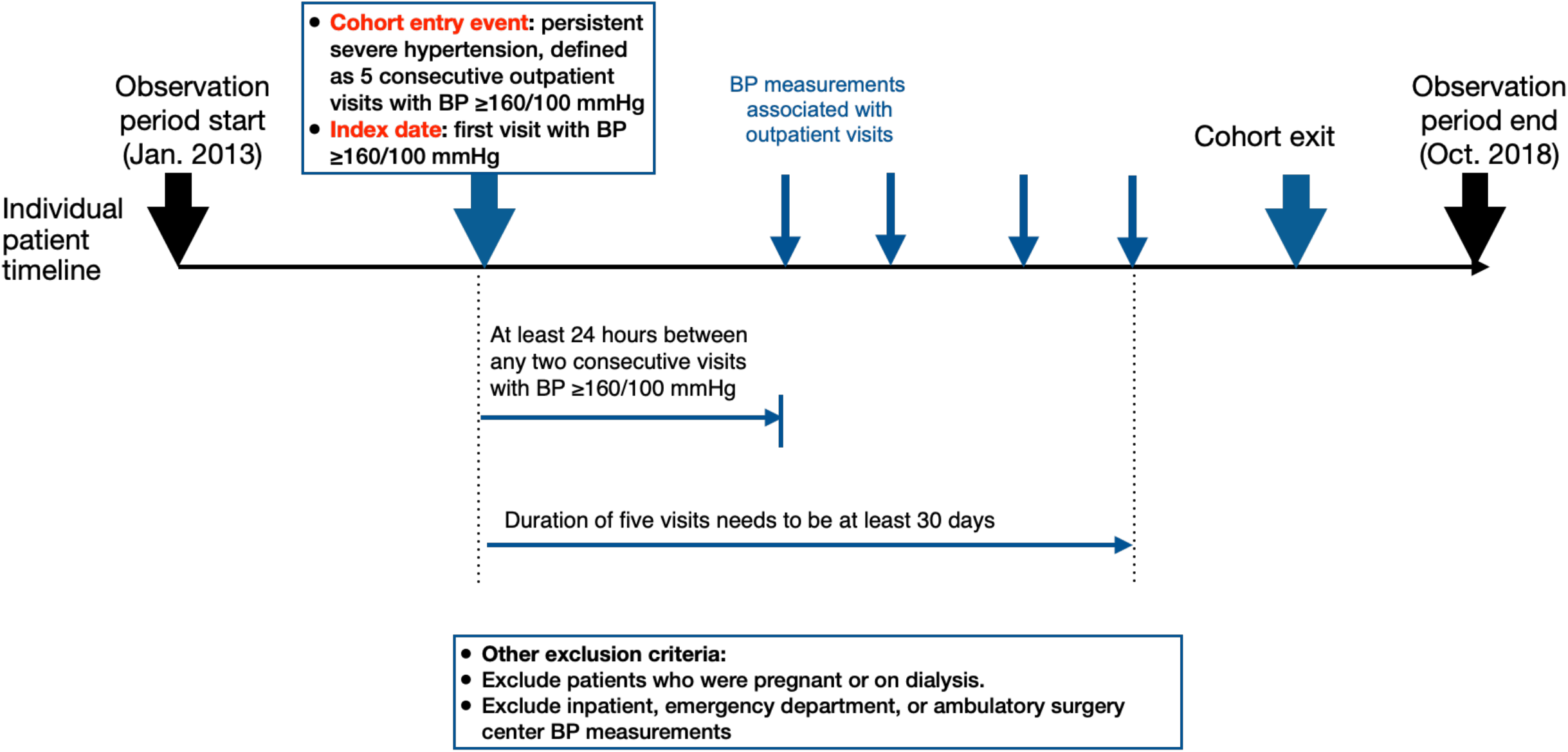
Cohort definition.

### Approach to Qualitative Analysis and Taxonomy Development

We used a systematic, inductive approach to develop a novel taxonomy of reasons for persistent hypertension based on available clinical data from patient medical records (**Figure 2**). This process has been previously applied to medical record review^13-17^ and was conducted in three steps as described below.

**Figure 2.**
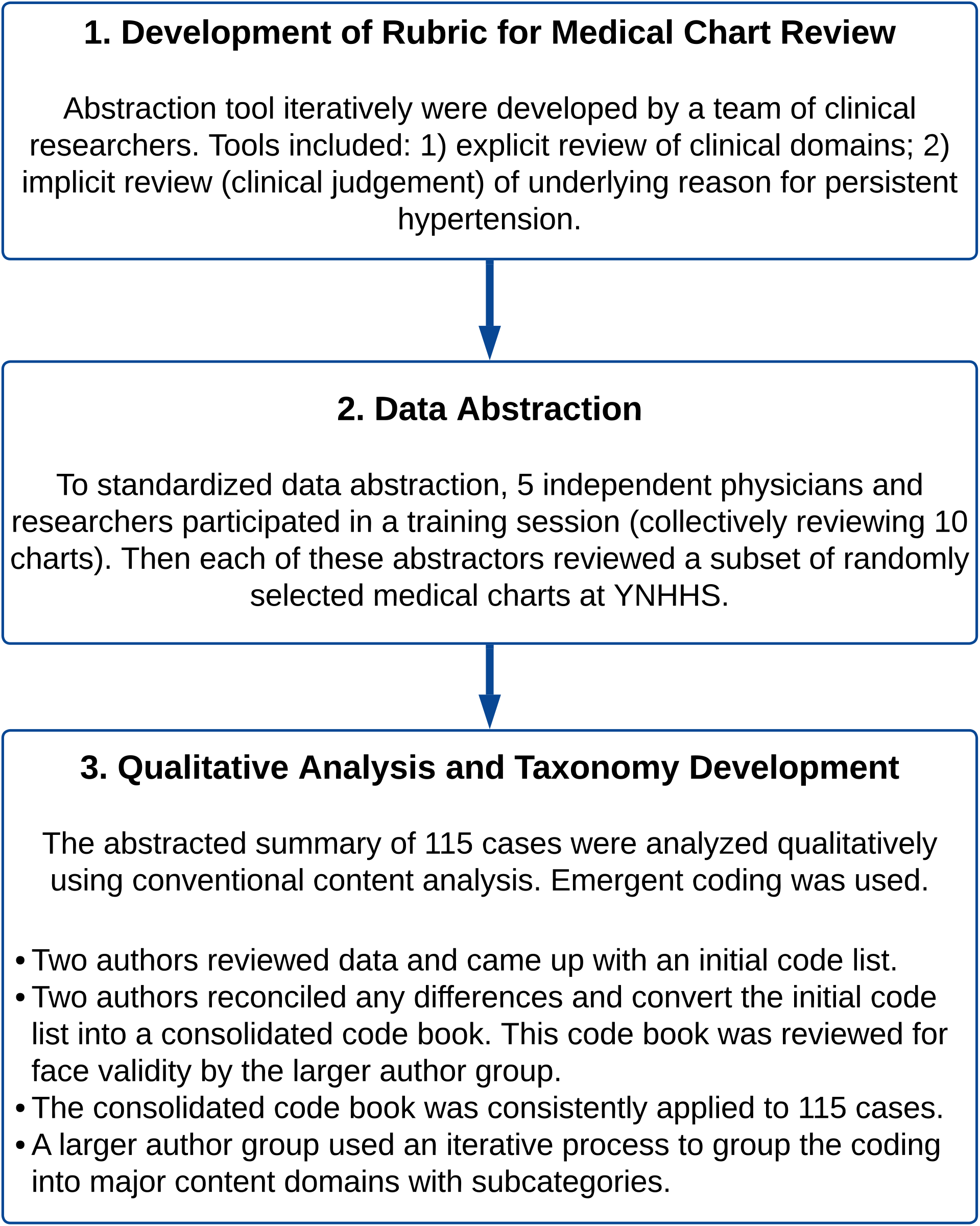
Three-step approach to taxonomy development.

#### Step 1: Development of Rubric for Medical Chart Review

Through an iterative process, a team of five clinicians and/or experienced cardiovascular researchers (Y.L., C.C., S.M., S.F, J.H) developed a rubric to systematically abstract data from the EHR (**Supplemental Table S1**). The team identified clinical domains relevant to the diagnosis and treatment of hypertension (including medical context of the encounter, BP measurements, medication prescriptions, medical history, laboratory results) and established criteria for consistency (to support explicit review). Additionally, the data extraction tool was designed to accommodate the clinical judgment of the reviewer on the underlying cause of persistent hypertension (implicit review). ^17-20^

#### Step 2: Data Abstraction

Five abstractors (YL, CC, SM, SF, JH) participated in a training session, during which they collectively abstracted 10 medical records using the rubric described above and generated a qualitative summary for each case, approximately 250 words in length. The abstractors reviewed both structured and unstructured data available in the medical records, including patient demographics, past medical histories, vital signs, outpatient medication prescriptions, laboratory results, encounter notes, and scanned documents. Decision rules and operational definitions were refined to reduce ambiguity and to facilitate standardized data abstraction. Discrepancies were resolved during face-to-face meetings with discussion among all reviewers until consensus was reached. Once the rubric was finalized, each abstractor reviewed a random sample of 20-30 medical records respectively. In the end, 115 of 200 cases were reviewed when reviewers determined they reached saturation; that is, no new constructs emerged from reviewing subsequent cases.^12^ Specifically, when reviewers felt they reach saturation, they reviewed another 10 charts to confirm no further constructs were identified.

#### Step 3: Qualitative Content Analysis and Taxonomy Development

The 115 cases abstracted using the rubric above were then further analyzed qualitatively using conventional content analysis. Content analysis is a systematic, replicable technique for compressing many words of text into fewer content categories based on explicit rules of coding.^9, 10^ Content analysis enables researchers to sift through large volumes of data with relative ease in a systematic fashion and it is useful in examining the patterns in documentation.^21^

We used emergent coding and established categories following preliminary examination of the abstracted data obtained in Step 2. First, two authors (CXD and YL) independently reviewed the abstracted data and developed a set of features to form the initial code list. Second, the two authors reconciled any differences that appeared up on their initial code lists. The initial code list was then developed into a consolidated code book (**Supplemental Table S2**). This code book was reviewed for face validity by the larger author group (YL, CXD, CC, HHK, HK, ES) and revised based on group discussion. Third, the consolidated code book was trialed on 10 cases by the coding group (CXD, YL, HHK) to ensure consistent application of the coding. The coding group checked that the reliability of the coding was established (agreement >95%, see details in **Supplemental Table S3**). We applied an inclusive coding strategy to mitigate the risk of assigning patients into a specific category. For example, if a patient has more than one issues with BP control, the coding group coded all issues for the patient. Then all 115 cases were coded by the coding group. Finally, a larger author group (YL, CXD, CC, HHK, HK, ES) used an iterative, consensus-based discussion process to group the coding into major content domains with subcategories, maintaining a consensus and primary data referencing approach.^9, 10^ We used NVIVO 12.0 (QSR International) software to assist with coding.

We described the demographic and clinical characteristics for the 115 patients included in the analysis. Comorbidities were defined using ICD-9-CM and ICD-10-CM codes based on the 1-year period prior to the index date (see details in **Supplemental Table S4**). We also described the mean (standard deviation [SD]) at the index date and across all visits, as well as the median (interquartile range [IQR]) time between visits.

## Results

### Study Sample Characteristics

We generated a randomized list of 200 patients and reached saturation with 115 patients. The mean age at the index visit was 66.0 (SD, 11.6) years; 54.8% were female; 52.2%, 30.4%, and 13.9% were noted in the EHR to be White, Black, and Latino/Hispanic, respectively (**Table 1**). A total of 25.2% had private insurance, 42.6% had Medicare, 20.9% had Medicaid and 6.1% did not have health insurance. The median (IQR) for total number of BP measurements used was 159 (140 to 177) for SBP, and 80 (71 to 90) for DBP. The mean (SD) SBP and DBP of the sample at the index date was 193.9 (13.9) mmHg and 92.5 (14.8) mmHg, respectively. The mean (SD) SBP and DBP of the sample across all visits were 158.8 (25.5) mmHg and 80.7 (14.6) mmHg, respectively. The median (IQR) time between visits was 11 (3 to 40). A large proportion of patients had comorbidities at the index date, including 41.1% patients with obesity (BMI≥ 30kg/m^2^), 22.6% with diabetes, 18.3% with chronic kidney disease, 26.1% with cancer.

**Table 1.**
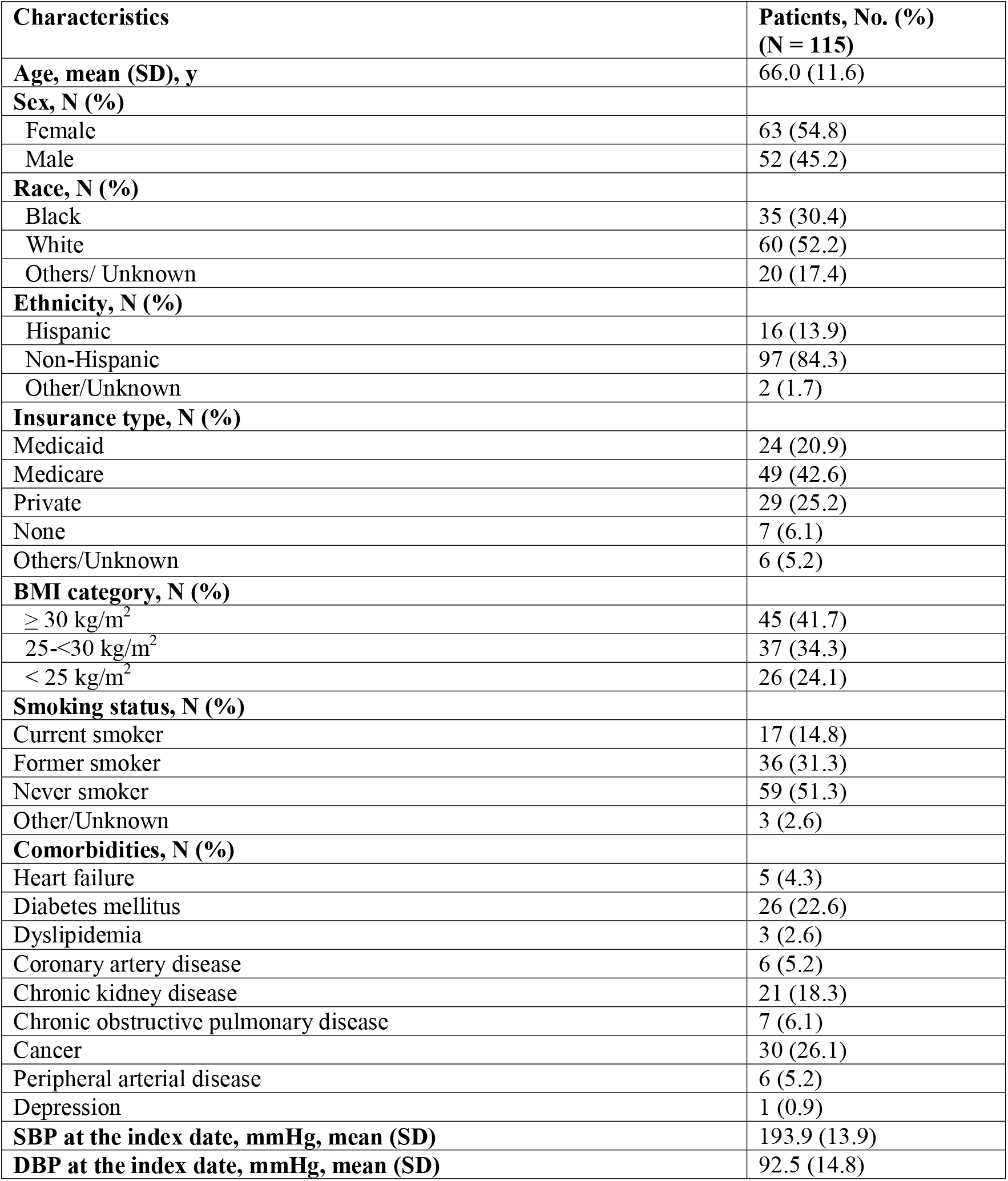
Demographics of 115 patients included in the sample for qualitative content analysis.

### Primary content domains

Based on content analysis of data available in the EHR for patients meeting our criteria, we identified a variety of subcategories grouped into three major content domains contributing to persistent elevated BP: non-intensification of pharmacological treatment, non-implementation of prescribed treatment, and non-response to prescribed treatment (**Tables 2-4**).

**Table 2.** Example quotations and clinical scenarios illustrating subcategories pertaining to non-intensification of pharmacological treatment

| Subcategory | Example quotation or clinical scenario |
| --- | --- |
| Provider purview | “Needs to see his primary MD for hypertension.” - Podiatrist |
| Competing medical priorities | “Hypertensive to 170s-180s systolic...Beta blocker contraindicated given bradycardia, Renin angiotensin aldosterone system blockade contraindicated given chronic kidney disease...would not add additional diuretic given diastolic heart failure has been very stable on current dose torsemide and might predispose to orthostasis.” |
| Patient preference | “Multiple office visits with BPs in 180s-200s... Patient is adamant she won't increase BP meds. She says she is already on a lot of medications.” |
| De-emphasis of the office measurement | “202/90...Hypertension - routine home BP monitoring shows excellent control. His BP is significantly elevated today, his home machine correlates well with my reading which is reassuring in terms of accuracy... He has otherwise had excellent control at home, and this may well be his documented white coat hypertension syndrome... If his BP stays elevated, he will call and we will discuss medication options.” |

#### Non-Intensification of Pharmacological Treatment

Non-intensification of pharmacological treatment was defined as lack of antihypertensive therapy intensification in response to persistent severely elevated BP. We identified four subcategories pertaining to non-intensification of pharmacological treatment, including provider purview, competing medical priorities, patient preference, and de-emphasis of office BP measurements (**Table 2**).

First, provider purview cases were coded as follows. These patients were medically complex and seen by multiple specialty physicians for different chief concerns, including concerns unrelated to hypertension. The office visits with elevated BP were with specialists who often do not treat hypertension, yet the BP was measured during the encounter. For example, office visits were frequently with orthopedics, podiatry, or radiation oncology whose providers do not routinely manage hypertension (example quotations are included in **Table 2**). Second, cases with competing medical priorities were coded as follows. These patients had competing medical priorities at a single encounter, for example, active cancer, chronic kidney disease, or congestive heart failure that impacted the options for hypertension treatment and had pressing symptoms or urgency that led providers to defer intensification of hypertension treatment. Third, at any given encounter, a provider would also have to weigh patient preference in the decision to alter the treatment plan. One subtheme that emerged around this was that patients had preferences against intensification of the treatment plan. Finally, cases with a decision to de-emphasis of office BP measurements were coded as follows. Escalation of antihypertensives was deferred because patients reported that the elevated clinic BP measurements were not consistent with their home measurements, where the decision to intensify treatment plan would then be deferred pending ambulatory BP monitoring.

#### Non-Implementation of Prescribed Treatment

Non-implementation of prescribed treatment was defined as a documentation of provider recommending a specified treatment plan to address hypertension but treatment plan not being implemented. We identified four subcategories pertaining to non-implementation of prescribed treatment, including issues with obtaining medications, psychosocial barriers, patient misunderstanding, and patient perception of a negative experience with the medication (**Table 3**).

**Table 3.**
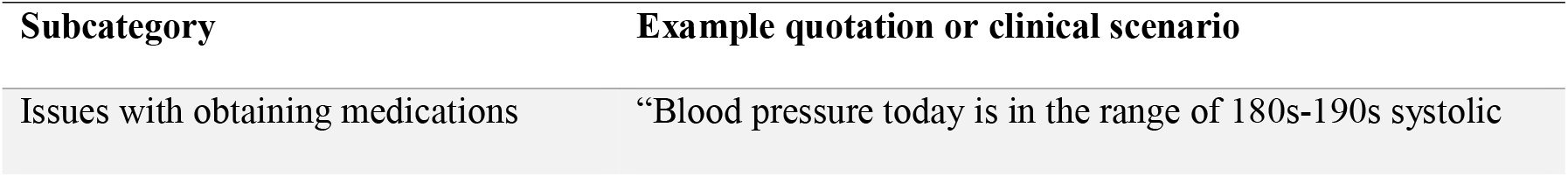
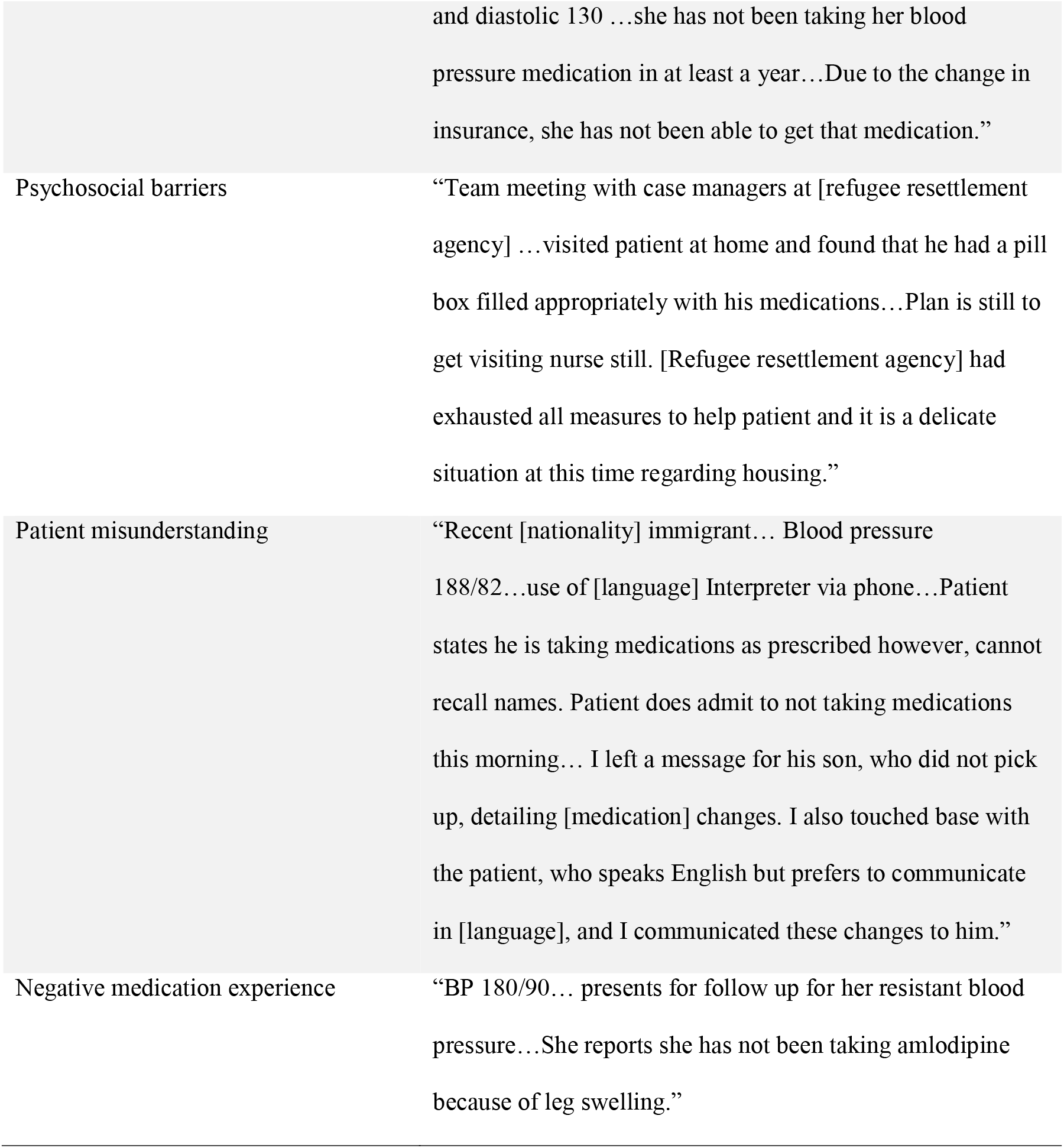
Example quotations and clinical scenarios illustrating subcategories pertaining to non-implementation of prescribed treatment

First, cases with issues of obtaining medications were coded as follows. These patients expressed that they could not follow treatment recommendations due to unmet health-related social needs, including socioeconomic barriers like unaffordability of medications or diagnostic tools for discerning treatment efficacy and lack of transportation to pick up prescriptions or pursue further diagnostic testing (example quotations are included in **Table 3**). Second, cases with psychosocial barriers were coded as follows. These patients had psychosocial barriers such as stressors around family conflicts, unstable housing, or employment that directly or indirectly prevented patients from taking medications as prescribed. Third, barriers such as insufficient education on the reasons for intervention from provider contributed to patient misunderstanding of the treatment plan, leading patients to be unclear on the treatment plan and thus not carrying out the plan correctly. Finally, cases with a negative experience with the medication were coded as follows. These patients stopped taking medications or implementing interventions due to the medication side-effects.

#### Non-response to Prescribed Treatment

Non-response to prescribed treatment was defined as clinician-acknowledged persistent hypertension despite documented effort to escalate existing pharmacologic agents and addition of additional pharmacologic agents and the presumption of patient adherence. We identified two subcategories pertaining to non-response to prescribed treatment, including resistant hypertension and secondary hypertension (**Table 4**).

**Table 4.**
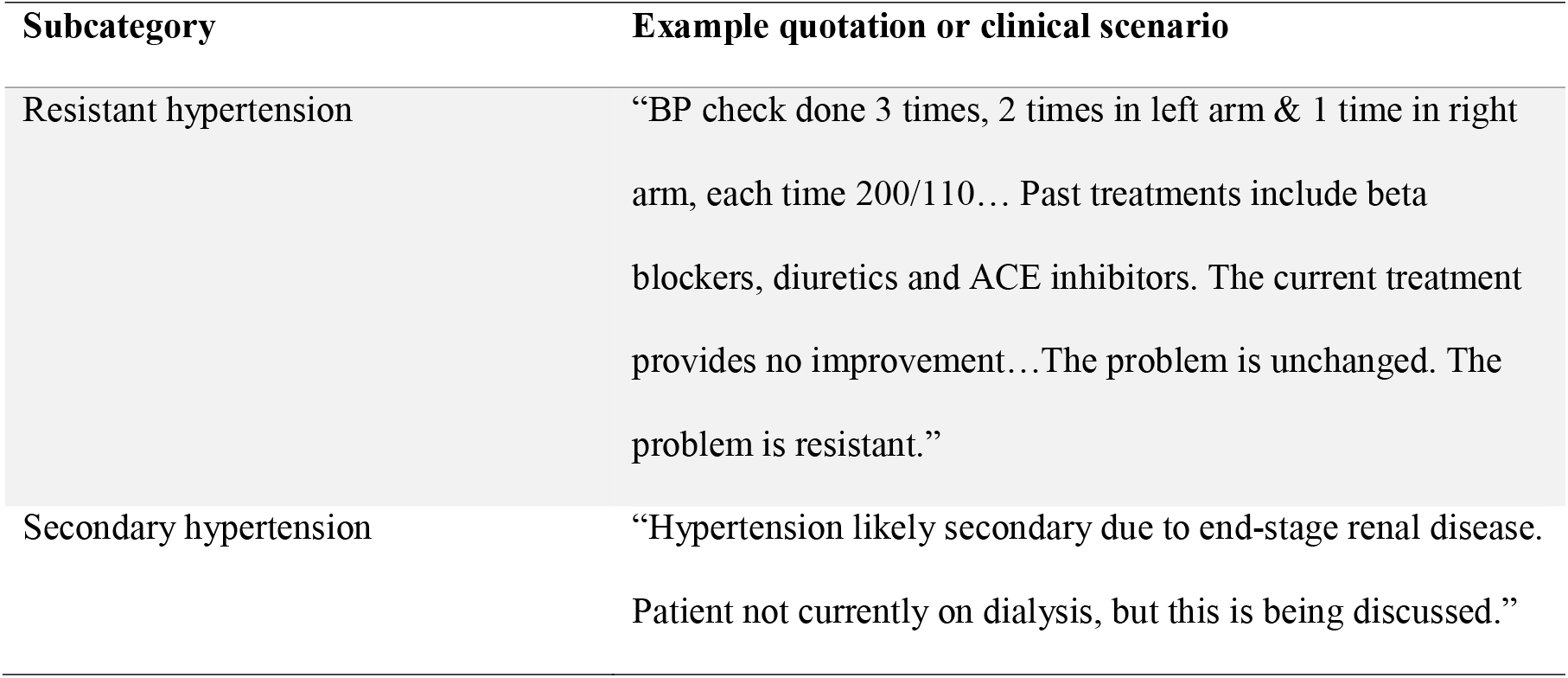
Example quotations and clinical scenarios illustrating subcategories pertaining to non-response to treatment

Resistant hypertension was defined as cases where the BP remains elevated despite concurrent use of three or more antihypertensive agents at maximumly tolerated doses.^6^ These patients were persistently hypertensive despite active treatment plan titration, including being prescribed three or more antihypertensive agents and having had secondary causes thoroughly investigated with negative results. Another subcategory identified was secondary causes of hypertension. These patients had a secondary cause of hypertension such as end-stage renal disease that was not immediately amenable to treatment.

#### Possible actions to address factors contributing to persistent hypertension

**Table 5** presents the different actions to address factors contributing to persistent hypertension. For example, to address provider purview cases, EHR integrated clinical decision support tools could send a notification to their primary care providers and automatically make a follow-up appointment. For patients who lack health insurance and have financial issues with obtaining medications, clinical decision support tools can help providers set up referral to community health workers to address health-related social needs. For patients with white coat hypertension, clinical decision support tools can facilitate providers ordering remote BP home monitoring for patients to obtain a more robust assessment of BP. For patients with misunderstanding of hypertension and medications, clinical decision support tools can help providers set up referral to patient education sessions. For patients with non-response to antihypertensive treatment, a smart orderset in the EHR can help providers set up referral to clinical specialists to further rule out secondary hypertension and titrate therapy with less commonly used medications and dosages. The taxonomy developed in this study can potentially be automated in the EHR and connected with EHR-based clinical decision support tools to improve care for patients with persistent hypertension.

**Table 5.** Possible actions to address factors contributing to persistent hypertension

| **Content domain** | **Subcategory** | **Possible actions to address the issue** |
| --- | --- | --- |
| Non-intensification of treatment | Provider purview | EHR notification to primary care provider's office to automatically make an appointment |
|  | Competing medical priorities | Discussion with patients for shared decision making |
|  | Patient preference | EHR referral to patient education program |
|  | De-emphasis of the office measurement | EHR smart orderset to assist physician to order home BP monitor |
| Non-implementation of treatment plan | Issues with obtaining medications | Screening for social risk factors; EHR referral to social worker |
|  | Psychosocial barriers | EHR referral to social worker |
|  | Patient misunderstanding | EHR referral to patient education program |
|  | Patient perception of a negative experience with the medication | EHR referral to patient education program |
| Non-response to treatment | Resistant hypertension | EHR referral to clinical specialists |
|  | Secondary hypertension | EHR Smart orderset to assist physician to order work-up for secondary hypertension |

## Discussion

This study presents a novel, pragmatic taxonomy for classifying patients with persistent hypertension by their contributing causes. Importantly, this actionable taxonomy is based on real-world data in the EHR, and it is likely that these classifications can be automated to provide better characterizations of the people who are failing to reach BP targets. This taxonomy was designed to act as a framework for developing EHR-based tools that could connect patients to targeted, high-quality care at scale.

Our study extends prior literature in several important ways. We introduced a conceptual framework to support the identification, classification, and intervention strategy matching for patients with severe hypertension who have persistent BP elevations over time despite contact with health care providers. The number of these patients is substantial, and their hypertension persists despite their obvious high risk and need for assistance.^22^ We previously suggested that a concept of persistent hypertension would better focus attention on the need to address the multiplicity of causes, of which resistant hypertension and its components are only a subset.^5^ Moreover, the persistence of hypertension for many individuals is a result of a failure in the implementation of antihypertensive strategies, which is a currently neglected facet of current classifications, particularly with respect to obstacles related to social determinants and structural racism.^23^ In this study, we further elucidated a variety of patient concerns leading to why a treatment plan was not carried out, including obstacles to obtaining medications, psychosocial barriers, patient misunderstanding, and negative medication experience, which further demonstrated the need for personalized social interventions, but also that these needs could be identified via information existing in the EHR. As such, this actionable taxonomy provides a more comprehensive approach to classifying people by the mechanism of what prevents them from reaching BP targets.

The key to the taxonomy is that different expertise and types of intervention are needed for specific categories and subcategories. For example, patients who lack health insurance and have financial issues with obtaining medications may benefit from referral to community health workers to address health-related social needs (**Table 5**).^24^ Patients with white coat hypertension may benefit from remote BP home monitoring to have a more robust assessment of BP.^25^ Patients with misunderstanding of hypertension and medications may benefit from patient education sessions.^26^ Patients with non-response to treatment may benefit from referral to clinical specialists to further rule out secondary hypertension and titrate therapy with less commonly used medications and dosages. For patients with persistently elevated BP at the orthopedist’s office, clinical decision support tools could send a notification to their primary care providers and facilitate setting up follow-up appointments.^27^ The BP measurements in almost all clinics at Yale New Haven Health System are performed using automated sphygmomanometers, so the measurements are relatively standardized. Although BP measurements may not be as accurate in certain specialty clinics if hypertension management is not their purview, it is unlikely that the measurement error would lead to a difference in BP value that is 20-30 mmHg higher than the BP control cutoffs (BP≥160/100 mmHg as defined in our study population). Our finding identifies a missed opportunity for helping these patients get the needed assistance to control their BP and prevent adverse outcomes. The bottom line is that a broad one-size-fits-all approach cannot address the multiple causes of persistent hypertension. Personalized medicine depends on distinguishing patients with persistent hypertension by their contributing factors; such knowledge is essential for tailoring care to individuals with appropriate evidence-based interventions.

Our findings have important clinical and public health implications. The actionable taxonomy developed in this study lays the foundation for developing effective tools for health systems to rapidly identify and classify people with persistent hypertension and connect them with targeted, personalized interventions at scale. One strategy is to use a learning health system approach built around a command center to promote population health.^28^ The command center can facilitate the screening of populations, identify high-risk population with persistently elevated BP, classify people by their underlying causes, match interventions with the specific issue that is impeding progress toward goals, monitor progress, detect spikes in risk meriting face-to-face assessment, and learn from the experience. The approach will be tested and iteratively refined in clinical practice to optimize analytics to determine risk and treatment response for individuals. It will also involve implementation science to determine how best to optimize the engagement of patients and improve their outcomes.^29^ This hub model will allow health care professionals to improve quality of care, optimize patient engagement and response, learn from experience, and improve outcomes across diverse populations. Essential to this approach is a technology backbone that enables the creation of an integrated and constantly updating database for each patient’s digital health data from all sources (e.g., health systems, laboratories, pharmacies, insurers, etc.). It is also essential to apply advanced analytics such as natural language process to distill information from unstructured clinical notes and machine learning methods to refine the algorithms for identifying high-risk patients. By leveraging digital data and digital technology, we have a great opportunity to accelerate the progress of hypertension control and meet the needs of patients and their providers.

This study has several limitations. First, we used an inductive approach to identify reasons for persistent hypertension using available information documented in the EHR. There may be systemic biases in what providers view as significant to document. For example, if there is no reason documented for a patient not taking a medication, it is unclear whether this was due to lack of clear documentation or to other system factors that make a provider not to explore the reason. Second, despite developing an iterative process for data abstraction and interpretation, the assignment of subcategories may be subjective. Third, this taxonomy was developed among a cohort of patients with persistent severe hypertension in a large US health system; it is not yet known whether the taxonomy will be applicable to other populations, for example, general patients with hypertension, whose contributing factors may also not fully be captured by the current classification system. Finally, the full value of this new taxonomy for classifying reasons for persistent hypertension will depend on whether it is used to inform future scientific investigations and clinical management aimed at improving the care and outcomes for these patients.

In conclusion, we propose a pragmatic, EHR-based taxonomy of factors contributing to persistent hypertension. This taxonomy can serve as a framework for developing EHR-based automatic tools that help classify patients with persistent hypertension and connect them to targeted interventions at scale.

## Data Availability

All data produced in the present work are contained in the manuscript.

## Non-standard Abbreviations and Acronyms

ABPM: Ambulatory blood pressure monitoring
BP: Blood pressure
DBP: Diastolic blood pressure
HER: Electronic health record
HBPM: Home blood pressure monitoring
SBP: Systolic blood pressure
SD: Standard deviation
YNHHS: Yale-New Haven Health System

## Acknowledgements

We thank Joshua Hillman and Soraya Fereydooni for conducting manual chart review of this study.

## Supplemental Materials

Tables S1-S4

