## Supplemental material for "Use of electronic health records to develop an actionable taxonomy of persistent hypertension"

Supplemental Table S1. Data abstraction form for manual chart review.

| **No.** | **Item** | **Example of phrasing** |
| --- | --- | --- |
| 1 | Age of the patient on 1^st^ visit | a black female in her 60s with history of diabetes, hypertension, and chronic lower back pain |
| 2 | Sex |  |
| 3 | Race/ethnicity |  |
| 4 | Comorbidities |  |
| 5 | 5 visits over what duration? | Had 5 visits of extremely elevated BP over 2 months |
| 6 | Seen by which specialist? | First 2 visits with IM, 3rd visit with physiatry, 4th and 5th visits with OBGYN |
| 7 | What medications was the person taking on 1st visit? What changes were made over the 5 visits? | At her 1st visit, patient was on Amlodipine but reported adverse effects to the medication. Amlodipine was stopped due to peripheral edema and patient was started on HCTZ 12.5 mg/day and asked to follow up in 1 month. On her 2^nd^ visit (4 weeks later), patient reported not taking any medications due to continuing concerns about adverse effects; doctor addressed concerns and patient agreed to start meds; asked to follow up in 1 month. 3rd visit was with physiatry where they noted that patient was still not taking her medications. 4th visit was at the OBGYN (2 weeks later), the patient called OBGYN for heavy vaginal bleeding, BP in the clinic was 171/68 mmHg, no intervention done only observed and told to f/u with PCP. 5th visit was at OBGYN where the patient reported having headache and was sent to the ED where her HCTZ was. Increased to 25 mg/d and she was asked to f/u with PCP in 1 week. |
| 8 | Any major lab tests done, evaluation for secondary hypertension, referrals made? |  |
| 9 | Any significant family history of hypertension? | Had family history of hypertension on both sides of parents |
| 10 | Any significant social determinants of health documented in notes (e.g., no insurance, need interpreter, homeless)? | Patient was concerned about the high cost of medication |
| 11 | Main impression for reason for persistently elevated blood pressure? | The main barrier to this patient’s BP control is non-compliance to medication |

*This was a hypothetical patient for training purpose.

**Supplemental Table S2.** Codebook used for qualitative content analysis

| **Higher Level Node** | **Sub-Node** | **Description** |
| --- | --- | --- |
| Did not take |  | Patient did not take prescribed medications/ interventions. |
|  | Could not afford | Patient could not afford medication/ intervention. |
|  | Could not obtain | Patient could not obtain medication/ intervention, due to reason other than affordability. |
|  | Intolerance | Patient stopped/ didn’t take medication due to adverse effect of antihypertensive medication. |
|  | Psychosocial barriers | Patient did not take medication due to psychosocial barriers, such as stress or depression. |
|  | Patient misunderstanding about intervention | Patient misunderstood or was uncertain on intervention/ med instructions, medication indication, benefit of medication contributing to not taking intervention (e.g., due to language barrier and health literacy). |
|  | No reason documented | Patient did not take medication, but no reason documented. |
| Did not intensify |  |  |
|  | Diffusion of responsibility | BP elevated at encounter with specialist that does not routinely manage hypertension, the specialist does not take responsibility for elevated BP and does not intensify treatment, and then follow up with a hypertension managing provider (i.e., PCP, cardiology, etc.) did not occur. |
|  | De-prioritized | Provider deferred BP med titration due to prioritization of other medical problems. |
|  | Patient preference | Provider did not intensify intervention due to patient preference not to. |
|  | Diagnostic uncertainty with BP measurement | Provider did not initiate or intensify treatment due to variation in BP measurements, either at home or in office, including cases where high in office readings were thought due to white coat effect. |
| Did not respond |  |  |
|  | Biologic non-response | Patient had elevated BP despite concurrent use of 3 or more antihypertensive agents at appropriate dose. |
|  | Secondary hypertension resistant to drug treatment | Patient had elevated BP due to another medical condition or suspected/ pending evaluation for another medical condition causing elevated BP and was not responsive to antihypertensive medication. |

**Supplemental Table S3.** Inter-rater agreement for 10 cases coded by the consolidated code book.

| Patient ID | Coder (listed by initial of reviewers) | | |
| --- | --- | --- | --- |
|  | CXD | YL | HHK |
| 1 | - Did not take: Intolerance - Did not intensify: Diagnostic uncertainty with BP measurement | - Did not take: Intolerance - Did not intensify: Diagnostic uncertainty with BP measurement | - Did not take: Intolerance - Did not intensify: Diagnostic uncertainty with BP measurement |
| 2 | - Did not take: No reason documented - Did not intensify: Diagnostic uncertainty with BP measurement | - Did not take: No reason documented - Did not intensify: Diagnostic uncertainty with BP measurement - Did not intensify: Diffusion of responsibility | - Did not take: No reason documented - Did not intensify: Diagnostic uncertainty with BP measurement - Did not intensify: Diffusion of responsibility |
| 3 | - Did not take: No reason documented | - Did not take: No reason documented | - Did not take: No reason documented |
| 4 | - Did not take: Could not afford | - Did not take: Could not afford | - Did not take: Could not afford |
| 5 | - Did not intensify: De-prioritized | - Did not intensify: De-prioritized | - Did not intensify: De-prioritized |
| 6 | - Did not intensify: Diffusion of responsibility | - Did not intensify: Diffusion of responsibility | - Did not intensify: Diffusion of responsibility |
| 7 | - Did not take: Could not afford - Did not take: Could not obtain | - Did not take: Could not afford - Did not take: Could not obtain | - Did not take: Could not afford |
| 8 | - Did not intensify: Diagnostic uncertainty with BP measurement - Did not take: Patient misunderstanding about intervention | - Did not intensify: Diagnostic uncertainty with BP measurement - Did not take: Patient misunderstanding about intervention | - Did not intensify: Diagnostic uncertainty with BP measurement - Did not take: Patient misunderstanding about intervention - Did not intensify: Patient preference |
| 9 | - Did not take: Could not afford | - Did not take: Could not afford | - Did not take: Could not afford |
| 10 | - Did not intensify: De-prioritized | - Did not intensify: De-prioritized | - Did not intensify: De-prioritized |

**Supplemental Table S4.** Diagnosis codes for comorbidities

| **Condition** | **ICD-9-CM** | **ICD-10-CM** |
| --- | --- | --- |
| Heart failure | 428 | I50 |
| Diabetes mellitus | 250 | E08, E09, E10, E11, E12, E13, E14 |
| Dyslipidemia | 272.0, 272.1, 272.2, 272.3, 272.4 | E78.0, E78.1, E78.2, E78.3, E78.4, E78.5 |
| Coronary artery disease | 414.0 | I25.1 |
| Chronic kidney disease | 585 | N18 |
| Chronic obstructive pulmonary disease | 490, 491, 492, 493, 494, 495, 496 | J40, J41, J42, J43, J44, J45, J46 |
| Peripheral arterial disease | 443 | I73 |
| Neoplasm | 140-239 | C00-D49 |
| Depression | 296,311 | F32, F33 |
